# COVID-19 Outbreak in Oman: Model-Driven Impact Analysis and Challenges

**DOI:** 10.1101/2020.04.02.20050666

**Authors:** Kashif Zia, Umar Farooq

## Abstract

Motivated by the rapid spread of COVID-19 all across the globe, we have performed simulations of a system dynamic epidemic spread model in different possible situations. The simulation, not only captures the model dynamic of the spread of the virus, but also, takes care of population and mobility data. The model is calibrated based on epidemic data and events specifically of Sultanate of Oman, which can easily be generalized. The simulation results are quite disturbing, indicating that, during a process of stringent social distancing and testing strategies, a small perturbation can lead to quite undesirable outcomes. The simulation results, although consistent in expected outcomes across changing parameters’ values, also indicate a substantial mismatch with real numbers. An analysis of what can be the reason of this mismatch is also performed. Within these contradictions, for Oman, regarding the eradication of epidemic, the future is not extremely alarming.

## 1 Introduction

COVID-19 is the latest evidence of epidemic disease capable of producing an extraordinarily large number of infections starting from a few [1]. According to Lippi and Plebani, Coronavirus disease 2019 (COVID-19), which originated in the city of Wuhan China on December 01, 2019, is a respirational and zoonotic disease, caused by a virus of the coronaviridae family [2]. The Virus strain is severe acute respiratory syndrome coronavirus 2 (SARS-CoV-2), resulting in fever, coughing, breathing difficulties, fatigue, and myalgia. It may transform into pneumonia of high intensity.

Towards successful diagnostic and cure of COVID-19, scientists in the field of molecular biology [3] are working hard to find answers about its spreading and infecting by examining virus samples. Although, the disease strain is known, but, the vaccine is no where near. And it is essential to ensure strict mitigation actions so that virus can be contained. Already, most of the countries are taking different kinds of precautionary measures to cope with it so that the losses can be reduced.

From the experiences of China and South Korea, the countries able to contain the spread of the disease so far, it is learnt that *social distancing* and *testing* are the key factors. Another factor responsible of spreading of the disease worldwide was regional and global travellers. Although, air travel is almost suspended now, but, this initial shock and countries (and people) not taking it too seriously has taken many countries in Europe and North America to a real bad situation.

In this overall scenario, after widespread suspension of air travels, Oman has taken various other measures to avoid spreading the infection. Schools and Colleges/Universities were closed from mid of March, followed by other non-essential offices and services. The situation is been monitored on daily basis by the government. However, there are many suspected cases which are not been tested so far. By the time of this writing, most of the country is partially locked downed. Although, the spread of the disease is not that much as of today (March 31, 2020). But, the new cases are appearing continuously. And the next few week are very important.

To know what may happen, we need to model and simulate. Towards this, we model the dynamics of spread of the disease (the epidemic model), population data and mobility of people. We have used a system that is designed to to it.

The epidemic model presented in this paper is not novel. In fact, similar (or even same) models are already proposed in different fields of study. However, contextualization and implementation of the model in the current global and regional situation is significantly important. Already such studies are been taken up by the research groups working in this area [4]. Through this paper, we have provided a focused analysis of the situation and asked important what-if questions, particularly in the context of Oman. However, the suggested method can be applied to any other country of the World, or even at the global level. Also, we have tried to argue why there is a big mismatch between real cases and cases predicted by the simulation.

## 2 COVID-19 Disease Spread Model

### 2.1 The Base Model

The model is based on well-established state-transition systems that are being used to study epidemics for a long time. The simplest one are SI (susceptible-infectious) and SIR (susceptible-infectious-recovered) models [5]. In both, an *infectious* individual infects a *susceptible* individual at a rate *β*. In SIR model, we also have a recovery rate (*µ*) after which an infectious individual is *recovered* permanently.

It can easily been seen that simple SIR model does not fully grasp COVID-19. For epidemics like corona-virus, SIR model was extended to SEIR [6], introducing a new state *exposed* between susceptible and infectious. This state is also known as *latent*, representing the period during which the individual has been infected but is not yet infectious himself. Therefore, we also have an exposed rate denoted by *ϵ*.

Still, the model needs further extension. The closet model representing the COVID-19 specifications is the one proposed for H1N1 epidemic [7]. Like in H1N1, in COVID-19, we have two types of infectious individuals; one that show symptoms (Symp) and the other who do not show symptoms (ASymp). And, an exposed individual can transit to state *infectious Symp* with rate *ϵ* or to a state *infectious ASymp* with rate 1 − *ϵ*.

### 2.2 The COVID-19 Model

The base model adapted from [7] is further extended to incorporate a new state *isolated* or quarantine. The isolated state represents the possibility of transferring an infectious individual with symptoms to isolation with a rate *α*. The value of *α* is then used to represent preparedness of health system of a country (or globe). All three states infectious Symp, infectious ASymp, and isolated, transits to recovered state with same rate *µ*. Also a transition from infectious ASymp to infectious Symp is made possible with a rate *ρ*. The final model is shown in Figure. 1.

**Figure 1:**
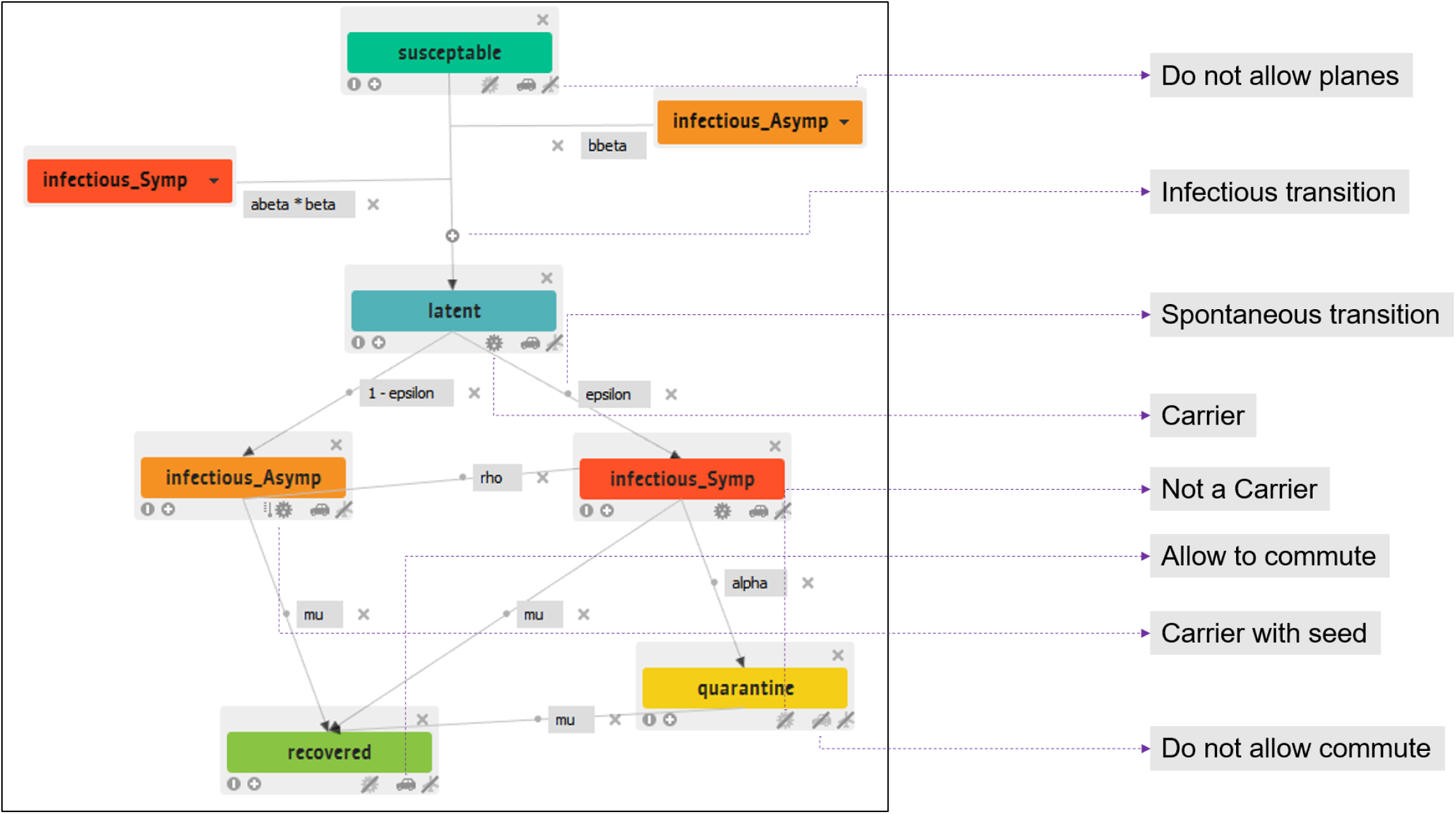
The COVID-19 system dynamics model.

## 3 Model Implementation

### 3.1 GLEAMviz Simulator

The model is implemented in GLEAMviz [8], the global epidemic and mobility model. GLEAMviz is a simulator which uses real-world data of population and mobility networks (both airways and commuting) on the server side. It integrates this data with the model developed by a user on the client side (similar to what we have presented in Figure. 1). Hence, the simulation generated in data driven, in which a user is responsible of describing system dynamics model of the epidemic, whereas, all the relevant population and mobility global data in integrated at the server side. As a result, the time-series data of spread of epidemics is generated by the server system.

In GLEAMviz client, the model is developed by showing transitions between different compartments (states). The model, conceptualized in the previous section, built on compartments and transitions is shown in Figure 1. One aspect not explained yet is that there are two type of transitions. The *infectious* transition is represented by “+” sign, depicting an addition of infectious cases. There is only one instance – from susceptible to exposed/latent – like that. The other transitions are *spontaneous* (represented by dot sign). The initial seed to the model is provided at compartment infectious Asymp, that is, few initial effectees as starting seed that do not possess any symptoms.

### 3.2 Model Specifications

The are two functional modes of the model.

#### 3.2.1 Carrier-ship and Mobility

An individual who is exposed to the virus (whether with symptoms or without) is the carrier of the virus. Therefore, a susceptible and recovered individual is not a carrier, while others are. Although, technically, an isolated/quarantined individual is a carrier, but, we have assumed that she/he is quarantined and is no longer able to transmit. Next are mobility possibilities. We have mimicked very recent situations to restrict or allow mobility. For example, considering all air traffic suspended, no compartment allows air travel. Whereas, all individuals who are not quarantined are allowed to commute locally. The commuting restrictions are further modified by using different transition variables.

#### 3.2.2 Transition Rates

The following are the transition rates from one compartment to another. Note, that there are quite a few refinement in the base model. Also, for a Greek alphabet, it’s symbol and text is used interchangeably. All rates vary between 0 and 1, inclusive.

- **beta**: infectious rate that transforms susceptible to exposed. This happens under the influence of both individuals with or without symptoms. To differentiate, we have taken **bbeta** as the *β* value for individual with no symptoms, and **abeta** ×*β* as the value for individual with symptoms. In this way, we are able to relate the infections incurred in different situations.
- **epsilon**: rate of transiting from exposed to infectious state. An exposed individual can transit to state infectious Symp with rate *ϵ* or to a state infectious ASymp with rate 1− *ϵ*. The value of *ϵ* is reciprocal of exposed period, which is equal to 5.2 days in our model [9].
- **rho**: rate of transiting from infectious ASymp to infectious Symp state. The value of *ρ* is reciprocal of symptoms appearing period, which is equal to 2.3 days in our model [9].
- **mu**: rate of transiting from being infectious or isolated to recovered. The value of *µ* is reciprocal of infectious period. It is taken 30 days in case of COVID-19.
- **alpha**: rate of transiting from infectious Symp to isolated state.

## 4 Parameterization and Cases

The variations in beta, abeta, bbeta, and alpha make up different cases corresponding to different situations. The other variables (epsilon, rho and mu) are kept constant. Variations are introduced systematically based on what is observed in the last one month and what are possible actions of the future. We have categorized different situations (cases) based on the outcomes, which are: extremely bad (case 1), extremely good (case 2), and intermediate (case 3).

### 4.1 Complete Inaction: Case 1

What can be worse than a complete inaction by the authorities? The default values assigned to the variables: *beta* = *bbeta* = 0.5, *abeta* = 1, and *alpha* = 0.001, were able to generate such a situation. In this case, *abeta* ×*beta* = 1.0 × 0.5 (infectious rate incurred by infected individuals with symptoms) and *bbeta* = 0.5 (infectious rate incurred by infected individuals with no symptoms) both are 0.5, depicting the basic setting with no differentiation. The fact that the rate of getting isolated in really low (*alpha* = 0.001), depicts that there is no effort yet put by the authorities to contain the epidemic. In the context of Oman, we have put a few cases in Muscat, Khasab, and Salalah as the starting cases which got symptoms, and ran the simulation for a year, starting from February, 26, 2020 (when a few such cases were reported). Even though the infection rate (*β*) is intermediate (only 1 out of 2 susceptibles are infected) and all flights are suspended (there are no outside influence), the results about spread of epidemic are really bad.

This is case 1 shown in Figure 2. It suggests that the outbreak would be rapid and extreme, reaching 0.2 million cases per day after 45 days of the outbreak, and then it would start dropping rapidly. 80% of the population would be affected. A comparison between real cases reported for the first 30 days and what the model generated (in case 1) is given in Figure. 4. There is a clear mismatch. One reason can be the lack of testing. As of today (March 31, 2020), Oman still have several thousands suspected cases (the cases without testing). But, the gap is till very high, indicating that there may be lack of data about Oman in the simulator database itself. Nevertheless, we are not going to see this case as reality after the actions been/being taken.

**Figure 2:**
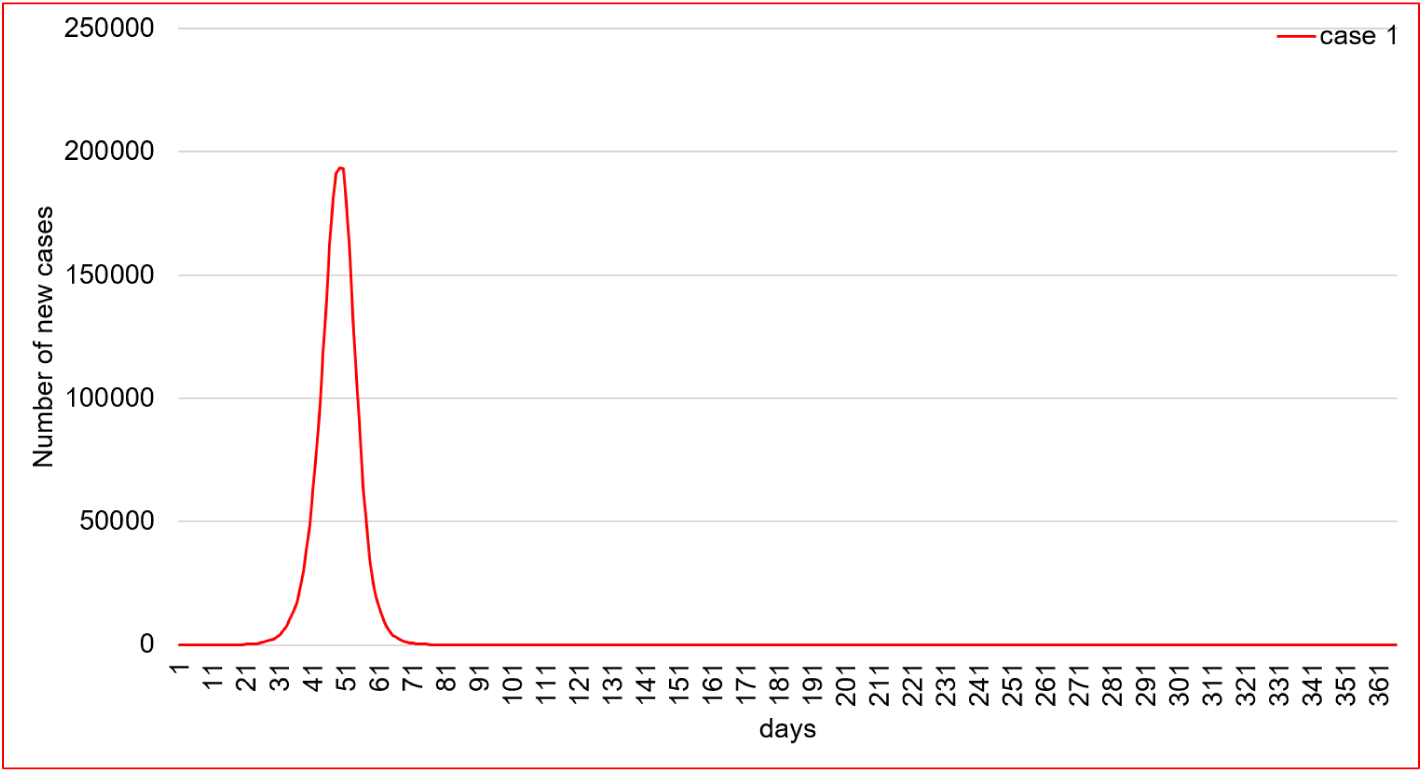
Infections (with symptoms): case 1

**Figure 3:**
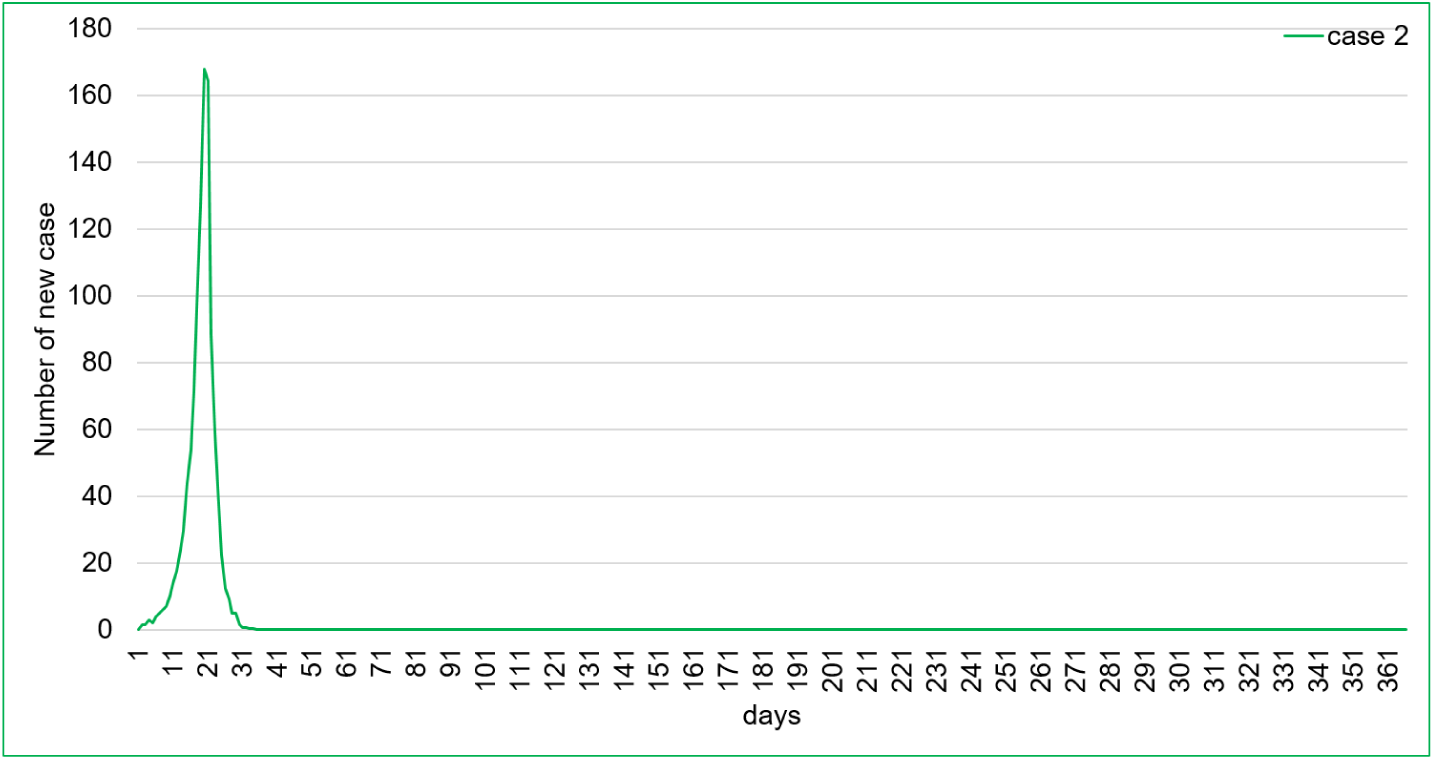
Infections (with symptoms): case 2

**Figure 4:**
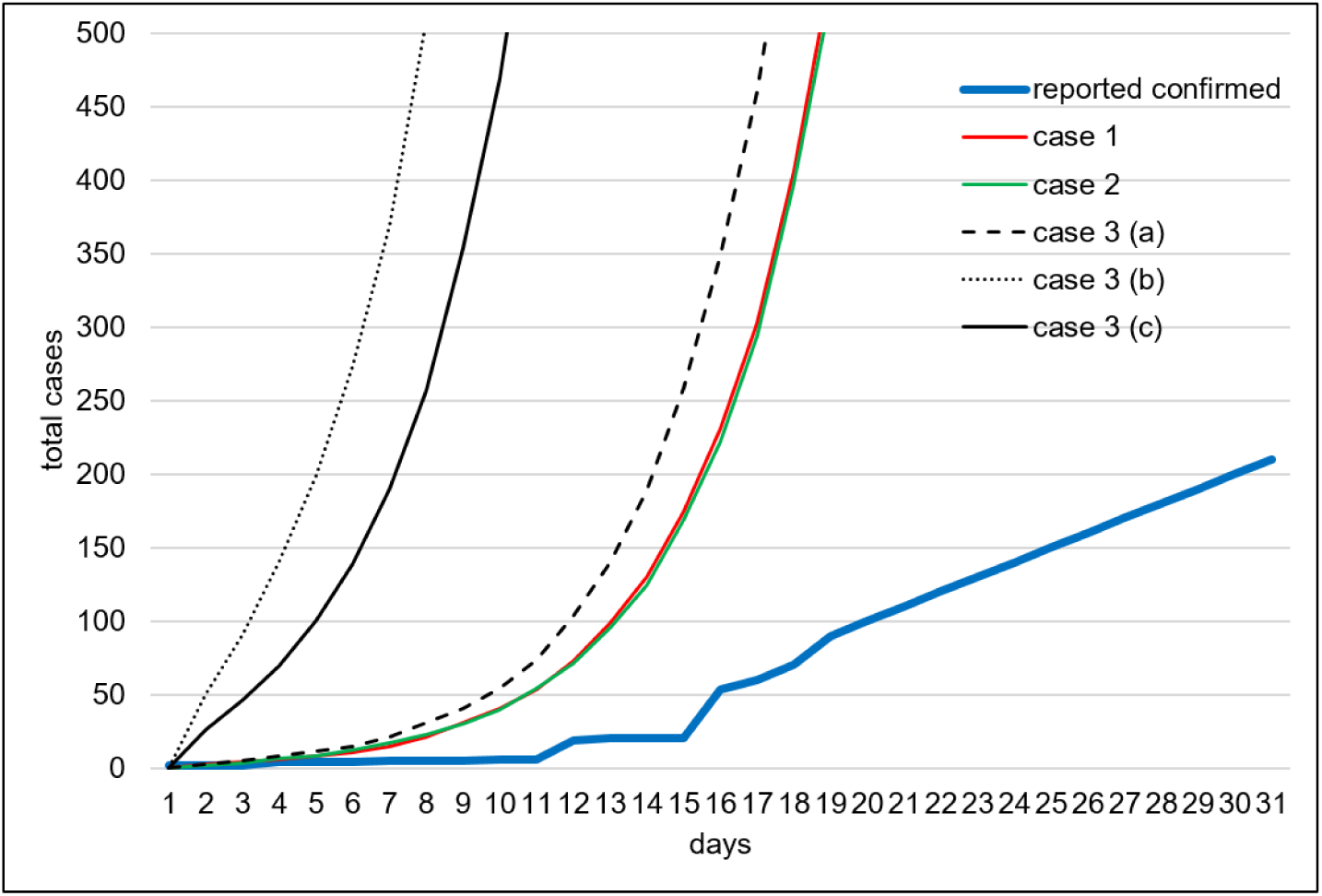
Infections (with symptoms) without suspected cases: comparison first 30 days

### 4.2 Extremely strict actions: Case 2

In this case, we try to mimic a *forced isolation* and a *limited lock-down* (enforced social distancing). This happens one week after the identification of the first cases in Oman. Many countries tried to put such restrictions for two weeks only, which were extended later on. We reproduced that by imposing if from March 3, 2020, but for 45 days. GLEAMviz provides an option to put exceptions in the form of rules to be applied for certain time period and for certain locations. The rules relate to setting the values based on mathematical expressions. For the above stated 45 days, we created exceptions, which are: abeta = 0.05, bbeta = 0.05, and alpha = 0.95.

What these values mean? The alpha = 0.95 means that infectious symp to isolated rate is 95% leaving only 5% patients to infect others. However, there is less probability of that happening due to lock down. Hence, we multiple abeta with beta to reduce it. bbeta is also reduced due to this reason. However, these reductions are quite strict and would be relaxed a bit in case 3. These exceptions were applied all across Oman. The curve shown in Figure 3 of case 2 simply shows that quite a few cases appeared in first week or so and then epidemic was eradicated.

Case 1 and 2, both are unrealistic, however, they educate us about extreme situations.

### 4.3 Realistic scenarios: Case 3

The real problem with this virus is that it may infect others from patients who do not have any symptoms. The authorities always report confirmed case. They are usually quarantined. But the threat is always there from people who are infected and in the society undetected. To represent this situation, we created the following exceptions:

- At the start of the simulation, introduced a few patients with symptoms and same number of patients without symptoms, at the three cities mentioned before.
- The following exceptions were applied (based on real events): after the first three weeks, real isolation activity was realized, first in small cities and after further three weeks all across Oman, by setting alpha = 0.95, and abeta = 0.05.
- A complete lock down was implemented after one month by setting bbeta = 0.05.

The above constituted an optimistic case named as case 3 (a).

Further, we introduced bulk cases (patients travelling and entering into Oman) into the three Omani cities.

Initially many of these cases were gone undetected. Avoiding any presumed extreme values, we opted to create two situations:

- case 3 (b): 20, 100, 20 patients
- case 3 (c): 10, 50, 10 patients

These people were integrated with their families and all of them were infected without any symptoms.

Many countries in the World are now aware that a lock-down of a few weeks would not be sufficient. Therefore, in this case, we also extended the lock-down indefinitely after first month. Hence, the results of this case can be considered as nearest to reality according to current actions of the authorities. Unfortunately, the results of the simulation were not good.

Starting with the optimistic case (case 3 (a)), we can see that the daily confirmed cases with symptoms reached to its peak of 3572 cases at day 42. Then they started to drop and reached to 0 at day 61. According to the timeline, day 42 is on April 7, 2020. In the context of the timeline, almost same pattern emerges for other two case (with bulk inclusions), but the numbers are much higher. Irrespective of the discrepancy between the reported cases and simulated ones, its a good news for Oman. We are close to peak and bracing for a downhill trajectory. Definitely, the containment policies should continue for at least one more month, hoping that epidemic does not start its second cycle.

Why case 3 (a) is closet to real data. According to the [10], on May 31, 2020, Oman had around 8000 suspected cases. If we just distribute this number equally after the first 10 days, and add them with the reported cases, we get a new curve (shown in Figure 5). This curve is very close to case 2 (a) curve, Actually it crosses it at day 28. There can be two reasons of this discrepancy. First is exactly the one we did the remedy for, that is, taking care of suspected cases. Second can be the lack of data in the simulator’s database itself. Simulator only has three cities of Oman. We did not notice much mobility across the populated coastal area. That can be one reason that the epidemic did prolong for only 60 days.

**Figure 5:**
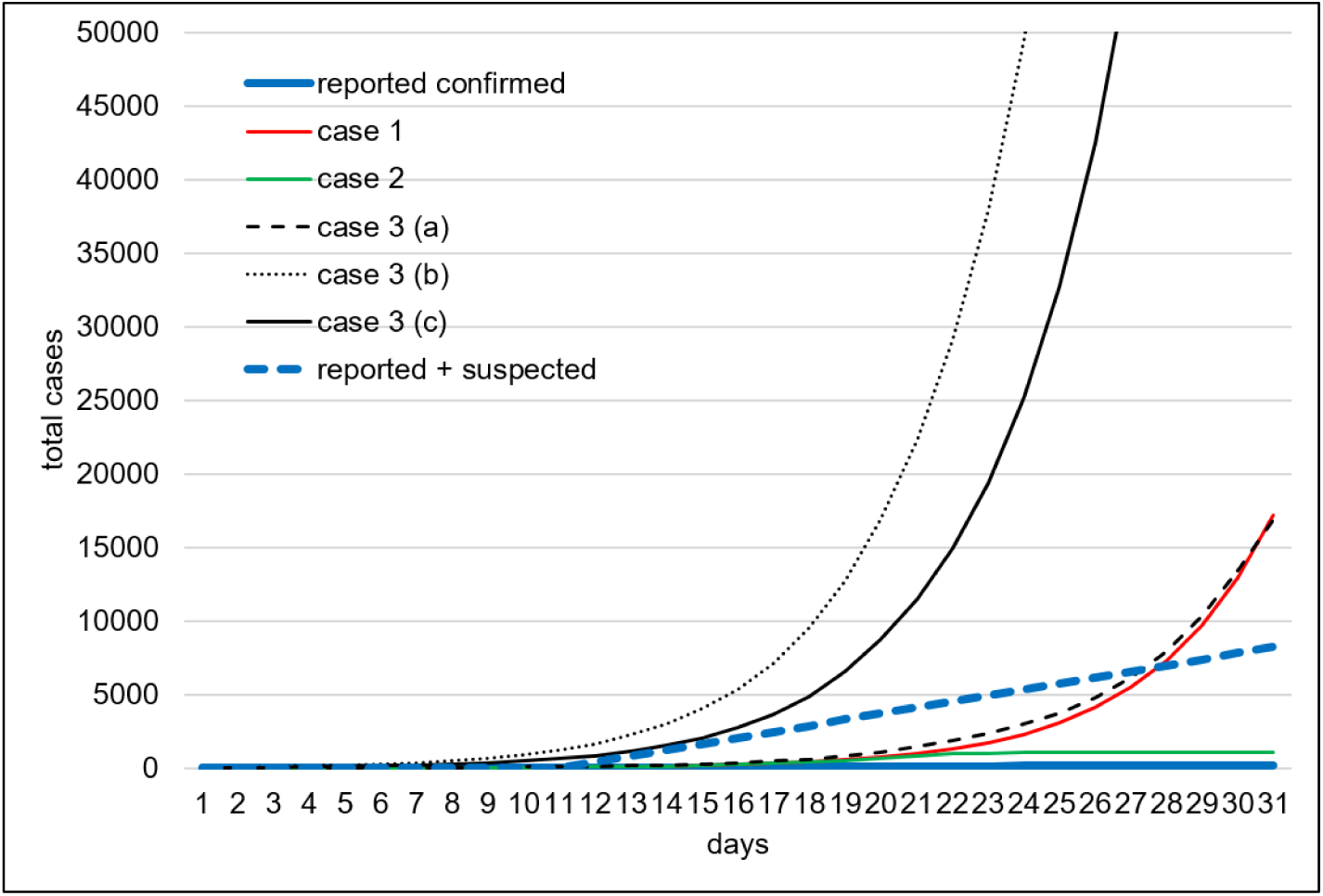
Infections (with symptoms) - with suspected cases: comparison first 30 days

**Figure 6:**
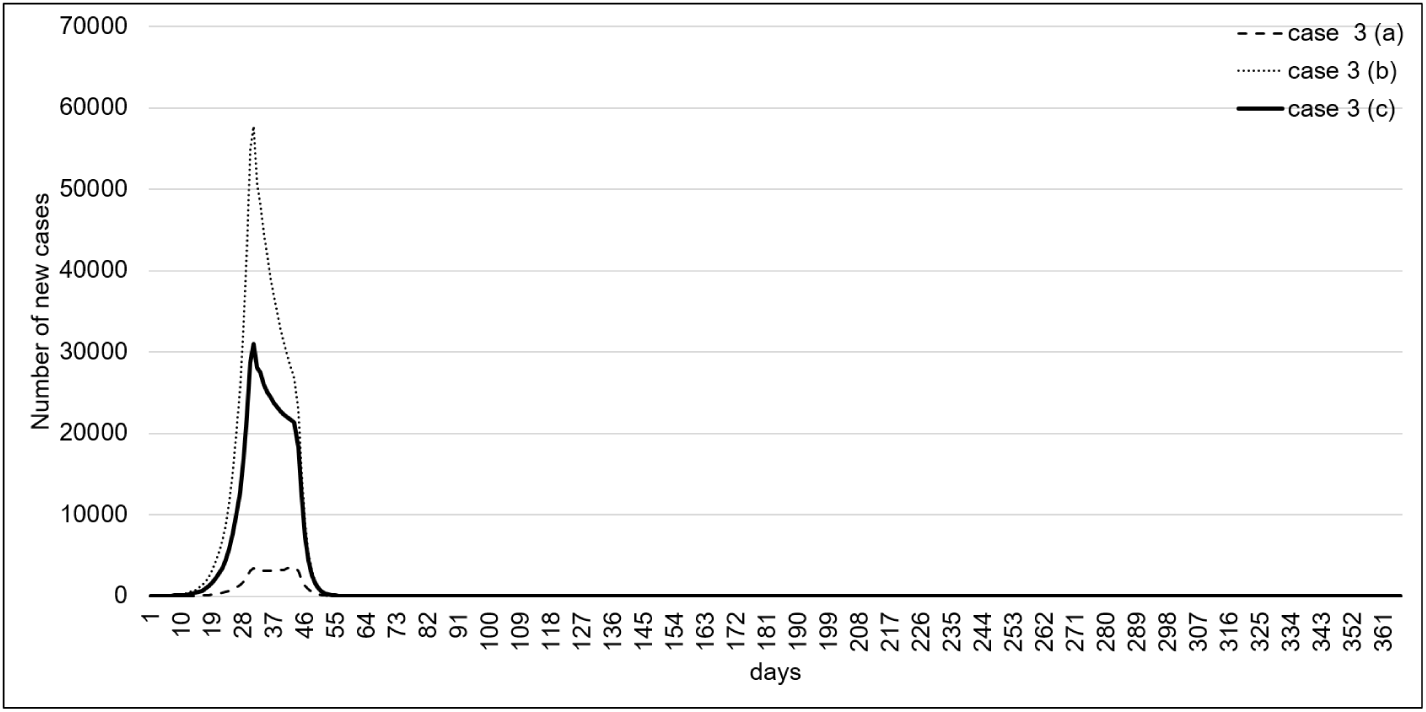
Infections (with symptoms): case 3

## 5 Discussion and Conclusion

With a system dynamic model of epidemic spread, incorporated with population and mobility data, we performed simulation of many different cases of COVID-19 impact, representing different real situations. The data used was only about Oman, but following the method adopted, the simulation can be performed for any country or region.

Like most countries around the World, Omani authorities took strict action against the spread of dangerous COVID-19 epidemic. Hence the first case (case 1) of unbounded spread, affecting almost 80% of population can easily be ignored.

The epidemic is becoming more and more dangerous, partially due to absence of strict actions early on. The actions required are social distancing and healthcare management (including testing and isolation of patients). A really strict action towards this is not possible as evidenced around the world. Hence the case 2 was also ignored.

In case 3, we implemented some real scenarios. We have seen that in case 3 (a), the epidemic’s peak is near, followed by a decline towards its eradication. However, we should be careful about it due to two reasons.

There is a substantial number of suspected cases, which have not neither been tested positive or negative. How many of these people are truly quarantined would be a deciding factor. The second aspect of it is inward traveling. As observed in China, a new wave of spread of epidemic may start due to people traveling into the country. Representing both these situations, as we can see in case 3 (b and c), just a dozen of these cases may have a big impact.

The simulator that was used has apparent limitations. There are only three cities for which the simulator has data. Also, during the running of the simulation, it was observed that mobility between the cities is almost non-existent. It is understandable, considering that cities of Muscat, Khasab, and Salalah are at the three corners of the country. With a more representative population and mobility data about different regions of the country, we may see more cases and a longer impact.

In fact, it was surprising to see that Oman has an epidemic timeline of only two months, which cannot even be imagined in current unfolding of the events. We performed a simulation with similar kind of settings for Pakistan and found out that the impact may extend to 8 to 9 months [11]. Nevertheless, the cases and results provide an educated guideline to the authorities about worst and best case scenarios and important factor leading to corresponding outcomes. The paper also guide other researchers towards modeling such systems and why they should be careful about putting forward predictions.

## Data Availability

No data that can be provided.

## Notes

### Competing Interest Statement

The authors have declared no competing interest.

### Funding Statement

No funding.

